# Exploring biomedical records through text mining-driven complex data visualisation

**DOI:** 10.1101/2021.03.27.21250248

**Authors:** Joao Pita Costa, Luka Stopar, Luis Rei, Besher Massri, Marko Grobelnik

**Affiliations:** Institute Jozef Stefan, Ljubljana, Slovenia

**Keywords:** Big Data, Semantic Technologies, Public Health, Healthcare, Text Mining, MeSH Headings, MEDLINE, PubMed, COVID-19, Diabetes, Mental Health

## Abstract

The recent events in health call for the prioritization of insightful and meaningful information retrieval from the fastly growing pool of biomedical knowledge. This information has its own challenges both in the data itself and in its appropriate representation, enhancing its usability by health professionals. In this paper we present a framework leveraging the MEDLINE dataset and its controlled vocabulary, the MeSH Headings, to annotate and explore health-related documents. The MEDijs system ingests and automatically annotates text documents, extending their legacy metadata with MeSH Headings. It then uses text mining algorithms that enable interactive data visualisations. These allow the user to the exploration of the enriched data made available by the MEDijs system.

**CCS CONCEPTS:** • **Information systems**; • **Computing methodologies → Machine learning approaches**;

**ACM Reference Format:** Joao Pita Costa, Luka Stopar, Luis Rei, Besher Massri, and Marko Grobelnik. 2018. Exploring biomedical records through text mining-driven complex data visualisation. In *Proceedings of SEBILAN ’21: ACM International Workshop on Semantics-enabled Biomedical Literature Analytics (SEBILAN ’21)*. ACM, New York, NY, USA, 6 pages. https://doi.org/0

## 1 INTRODUCTION

The importance of evidence in decision-making in public health and healthcare are relying today in the text mining and machine learning technologies that have been entering the health domain at a slow and cautious pace. The emergencies caused by the recent pandemics and the need to act fast and accurate were a motivation to fast forward some of the modernization of public health and healthcare information systems. Though, the amount of available information and its heterogeneity creates obstacles in its usage in meaningful ways.

The proposed MEDijs framework aims to facilitate the health professionals and researchers in the exploration of their own data, independently of their technical capabilities. Furthermore, it builds on well-established tools that are well known to health professionals (such as, e.g., the PubMed biomedical search engine and its open dataset MEDLINE) to offer an incremental level of difficulty in the usage of the framework, not compromising its usefulness in the health domain.

Since its declaration in March 2020 [18], the pandemic situation in Europe and arriving to the USA motivated the multiplication of available COVID-19-focused platforms (e.g. [20] or [10]), competitions (e.g. [13]) and open resources (e.g. [16]). This is an example of the current trend arriving also to the health domain make available healthcare information and seek for useful insights on that data that can lead to, e.g., new biomarker identification or evidence of the impact of other diseases when in relation to the new coronavirus. This is much motivated by the coordinated effort to fight this pandemic globally, in which part of this work is a contribution to [11].

## 2 THE MEDIJS FRAMEWORK

It is well known that, in particular in the scientific domain, the appropriate query can lead to a good hypothesis and is half way to the solution. Citing the American medical researcher and virologist Jonas Salk, “What people think of as the moment of discovery is really the discovery of the question.” Nowadays, the huge amount of data available can be a challenge to get the meaningful information needed, and the health domain is no exception. The well-structured open data and the smart systems that make the appropriate use of it are valuable and can help health researchers and professionals asking the right questions. That is the aim of the proposed approach in this paper.

The MEDijs workflow begins with the data ingestion, preprocessing and annotation of health-related documents, enriching them with MeSH headings based on their content. We will discuss this MeSH-based text classifier later in this paper, which performs the assignment of the MeSH Heading classes that will allow some of the insightful interactive data visualisation, as well as the integration in other systems (e.g. a news engine with a similar workflow to PubMed, also discussed later in this paper). The ingested documents can be of all sorts, from electronic health records to medical reports. They need to be written in english language, which is the base language of the controlled vocabulary MeSH and the dataset MEDLINE that we are using to learn our algorithms. At the moment we haven’t explored the possibility to include other languages, although we are aware of other multilingual approaches (such as [3] and [2]).

In the Figure 2 we present the architecture of the MEDijs system that implements the framework proposed in this paper. The ingestion of the most recent MEDLINE dataset and corresponding MeSH controlled vocabulary is fundamental to the basic use of the MEDijs system, as it serves as base to the machine learning algorithms that are used to automatically annotate the input text. The metadata of the input datasets ingested by the user will then be enriched with the MeSH annotation (that will will explain in detail later in the paper) and stored in the MEDijs database. The latter is based on the elasticsearch technology [17], allowing for powerful Lucene-based queries that will be used to enable the data visualisations ahead. Those queries can be used to explore the ingested and enriched data in a meaningful way.

**Figure 1:**
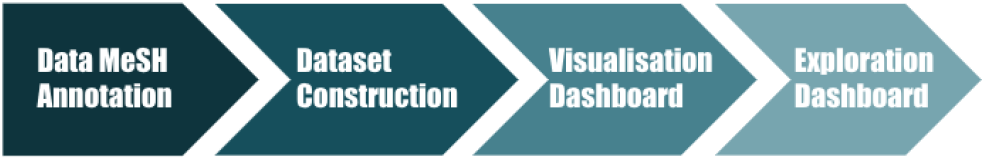
A workflow to explore biomedical documents.

**Figure 2:**
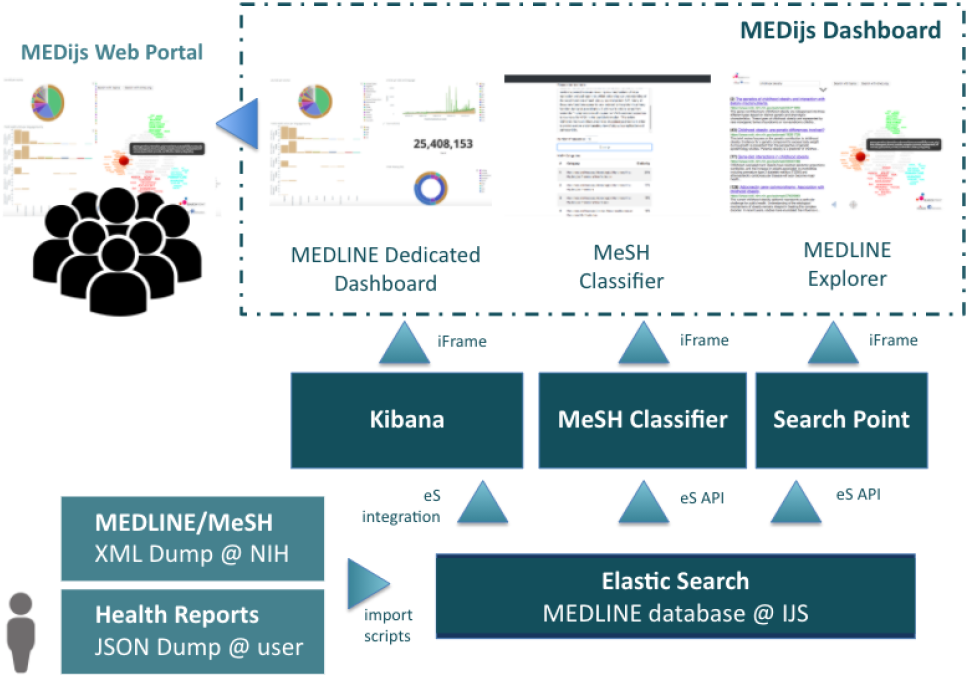
The MEDijs architecture and data exploration platform.

Using the elasticsearch queries we feed our text mining algorithms to enable interactive data visualisation modules. That is done first through the availability of on-demand data visualisation modules that can integrate several topic-focused dashboards (e.g. diabetes or mental health). These are built by the user based on saved samples of queried data and can be easily manipulated by the user, independently of his/her technical skills. This part of the system allows to explore both the data ingested, with its metadata enriched by the MeSH headings classification, and the MEDLINE data itself limited to the selected fields at the time of its ingestion.

The MEDLINE explorer that we make available allows the user to investigate the ingested data based on the Lucene-based queries using the metadata or key-phrases. It provides the user with a cluster of subtopics that relate to the query and a movable target over them that will reposition the obtained results in order to refine the search. This tool is of great usefulness to medical research.

We also made available a web-portal where the user can access the MeSH classifier directly, by dragging and dropping snippets of text to be annotated. This allows the user to interact directly with the MeSH classifier, and explore its potential in the annotation of health-related documents. We will be discussing this further in the following sections.

This framework is implemented through a web portal (located at www.qmidas.eu) where the anonymous visitor can experiment the MeSH classifier, the MEDLINE data explorer, and with an awarded password can also access the interactive visualisation dashboards and on-demand visualisation builders. It also allows for its integration through iframe or REST API, particularly in what respects the access to elasticsearch queries, the automatic annotation of text snippets with MeSH headings, and in the interaction with the MEDLINE explorer available through MEDisj.

## 3 THE CONSTRUCTION O THE DATASET

In 2020 the MEDLINE dataset [15] contains more than 30 million citations and abstracts of the biomedical literature dating back to 1966. Over the past ten years, an average of a million articles were added each year. Around 5% of MEDLINE is on published research on infections, with cancer research being the most prevalent occuping 12% of this body of knowledge. Most scientific articles in this dataset are hand-annotated by health experts using 16 major categories and a maximum of 13 levels of deepness. The labeled articles are hand-annotated by humans based on their main and complementary topics, and on the chemical substances that they relate to. It is widely used by the biomedical research community through the well-accepted search engine PubMed [19]. The richness of this data allows us for insight on aspects of the disease as well as approaches and best practices from the published research.

The accuracy of health vocabulary and selected concepts is fundamental to appropriately identify the information that needs to be extracted. Though, it is not always the case that health-related documents follow a concise vocabulary, which can be the cause for misleading summarized information and query results. Aiming to address this necessity, we have developed a text classifier [12] that learns over the hand annotations of MeSH Headings of biomedical research papers in MEDLINE, to provide the automated annotation of health-related text. The classification of the input text will provide added information that is more accurate than the key-phrases-based queries and improve the understanding of different types of health documents, from news articles to medical reports. We will be analysing some of these applications towards the end of this paper.

To have an efficient automated annotation of free text based on the already existing health-related categories provided by the MeSH Headings, we use the nearest centroid classifier [14] constructed from the abstracts from the MEDLINE dataset and their associated MeSH headings. Each document is embedded in a vector space as a feature vector of TF-IDF weights (where those weights are statistic objects that reflect how important a word is to a document in the collection). For each category, a centroid is computed by averaging the embeddings of all the documents in that category. For higher levels of the MeSH structure, we also include all the documents from descendant nodes when computing the centroid. To classify a document, the classifier first computes its embedding and then assigns the document to one or more categories whose centroids are most similar to the document’s embedding. We measure the similarity as the cosine of the angle between the embeddings. Preliminary analysis shows promising results. The classifier gives an order of positioning of all the MeSH categories annotating the provided free text.

The demonstrator version of the MeSH classifier is available online through a web portal (represented in Figure 3) provides the position number in the annotation and the percentage representing the weight of the MeSH term in the annotation (given by cosine similarity). The classifier is also available through a REST API, using a POST call, and taking a JSON input that includes the text input. We evaluated the classifier by leaving out one year of MEDLINE abstracts to be used as evaluation dataset. For this reason, the classifier has been trained on the MEDLINE 2018 dataset (27.837.540 article abstracts), leaving the latest batch of annotated abstracts out. Then, the model has been evaluated against new data from the MEDLINE 2019 dataset (325.128 articles). The main goal was to determine the performance of the MeSH classifier for medical texts. Additionally, the evaluation results should provide an estimate for an optimal similarity cut-off, classification depth and a decision regarding the classification of all MeSH terms or major classes only. We evaluated several combinations of the three parameters: similarity cut-off 15% to 50%, MeSH three depth from 1 to 7, and the usage of major classes only vs. usingall MeSH classes. For those combinations we have calculated the average precision, recall, F1 and F0.5 measures and concluded that the classification of all MeSH classes performs significantly better at the desired depth of classification, depth three. At that classification depth 3 it is estimated that the optimal similarity cut-off is around 0.25, providing F1=0.64. These parameters are then used to further optimise the classifier and evaluate it for the classification of health documents.

**Figure 3:**
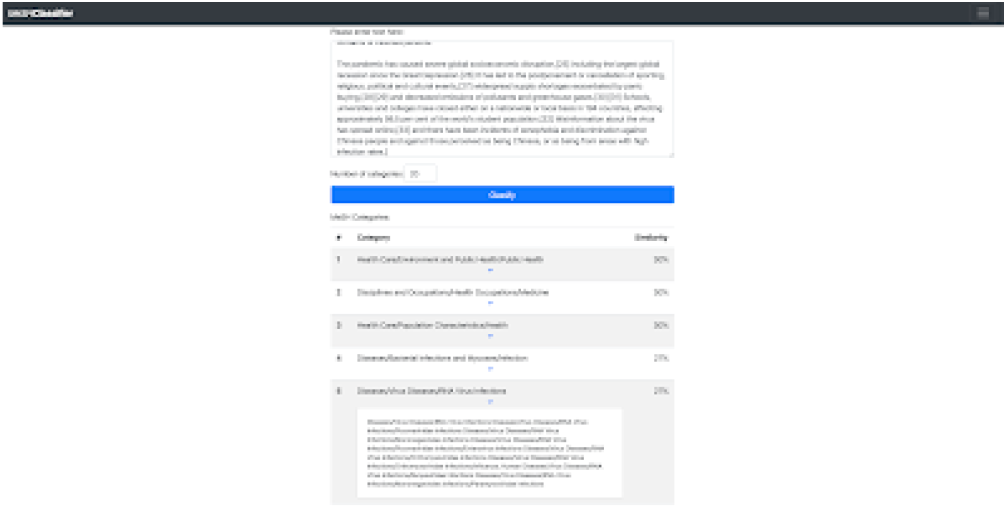
The MEDijs MeSH classifier for health-related documents.

**Figure 4:**
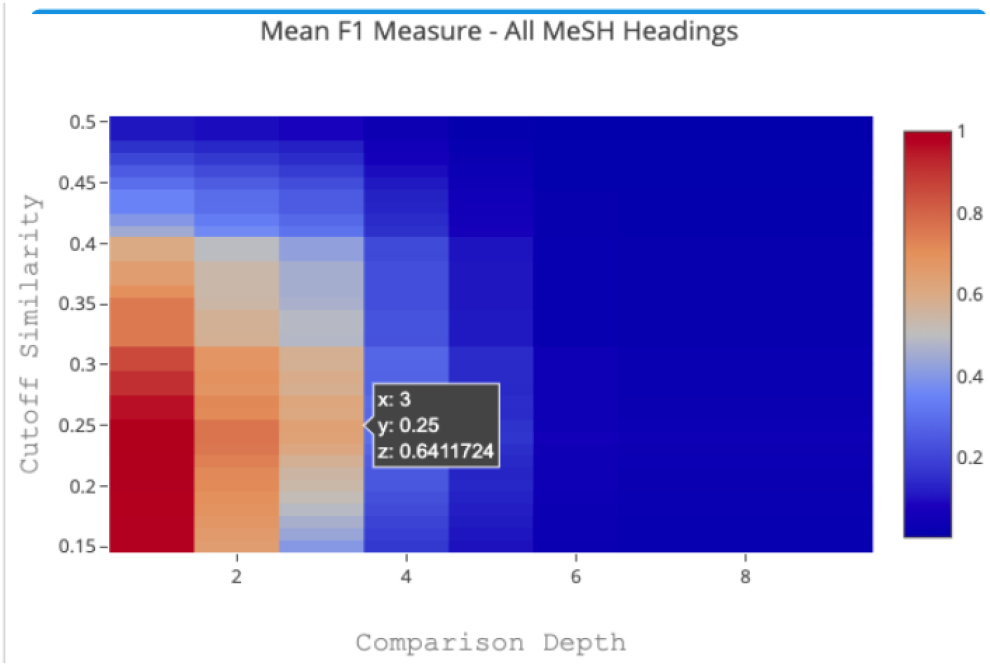
The evaluation of the MEDijs MeSH classifier.

## 4 ON-DEMAND DATA VISUALISATION

To guarantee some independence to the less technical staff we have developed a data visualization dashboard that provides the user with real-time access to a local data set sourced on the national volunteer participants. This is based on Elastic Search technology, together with the Kibana open source data visualization plugin. Part of this work was developed in the context of the European Union research project MIDAS [5], by applying the know-how obtained in building a similar system to monitor and manage the scientific knowledge open data set MEDLINE [15].

The input data enriched by the automated classifier, is delivered to the dashboard through an API to the main platform. The update of the back-end system is driven by import scripts that appropriately load the new dataset into a new index in Elastic Search. This new input data index generates the database that serves the system. The dedicated dashboard based on Kibana has a native integration with Elastic Search and, therefore gets the index imported automatically to dynamically build the new visualization modules and dashboards. The public instance that can be derived from a dashboard is dependent of the choices in the definition of that dashboard.

In the example in Figure 5) we can see the three most relevant views of this exploratory system. The first one - discover - allows the user to explore the ingested data and associated metadata. (S)he can later save the data sample queried to pass on to build the data visualisation modules from templates - Visualize. Then, the user can take those data visualisation modules and compose topic-focused dashboards populated with interactive charts and heatmaps that can provide insight to the user and be shared as public instances.

**Figure 5:**
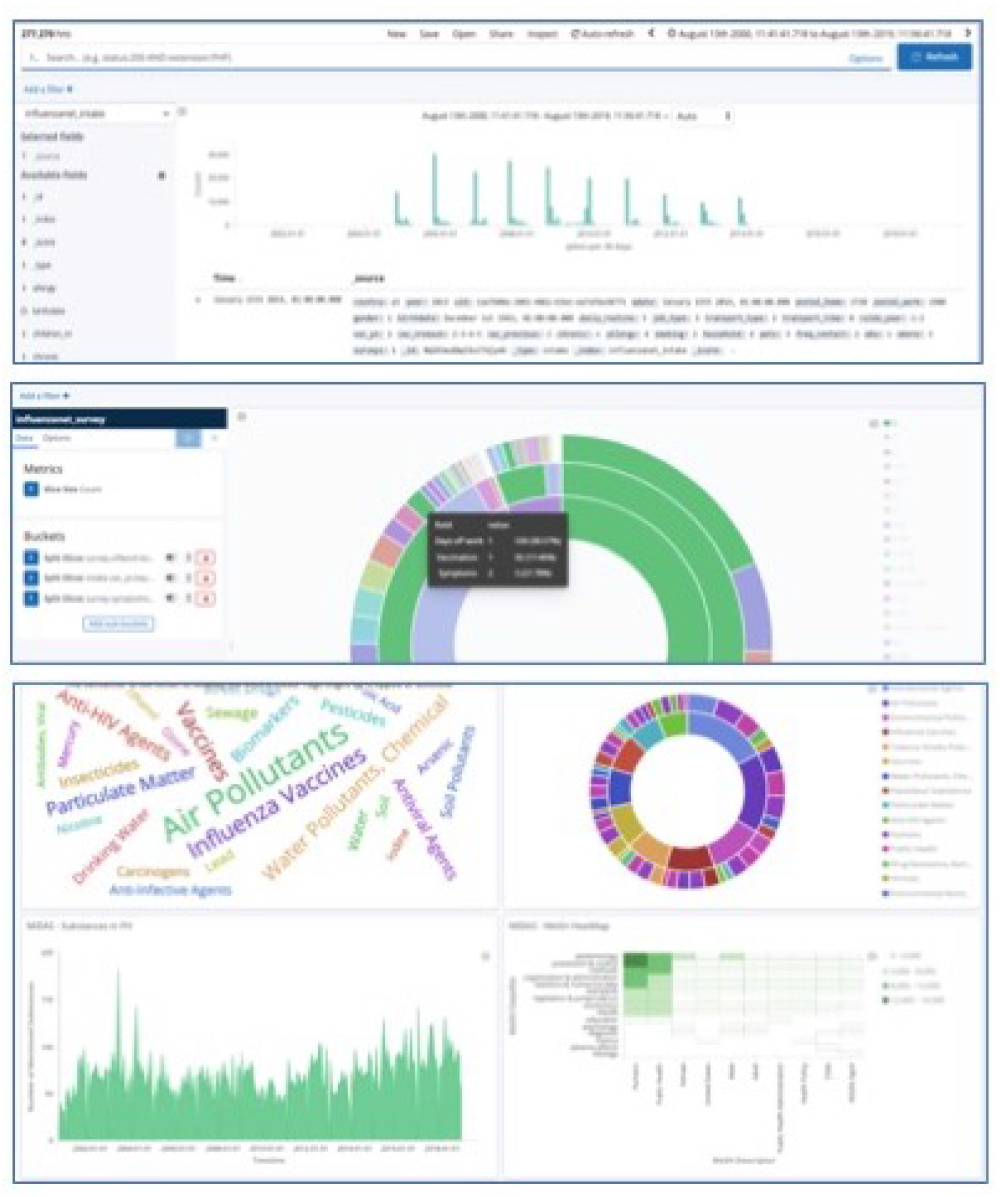
The data exploration tool allowing aiming for prototyping by health experts.

## 5 MEANINGFUL VISUAL EXPLORATIONS

To better understand the disease, the published biomedical science is the source that provides accurate and validated information. Taking into consideration a large amount of published science and the obstacles to access scientific information, we made available a MEDLINE explorer where the user can query the system and interact with a pointer to specify the search results (e.g., obtaining results on biomarkers when searching for articles hand-annotated with the MeSH class “Coronavirus”).

To allow for the exploration of any health-related texts (such as scientific reports or news) we developed an automated classifier [8] that assigns to the input text the MeSH classes it relates to. The annotated text is then stored in Elasticsearch [17], from where it can be accessed through Lucene language queries, visualized over easy-to-build dashboards, and connected through an API to the earlier described explorer. The interactive MEDLINE data exploration, referesented in the screenshot of Figure 6 shows a scientific article originally positioned as 182nd, now in the 4th place. (read [11] for more detail).

**Figure 6:**
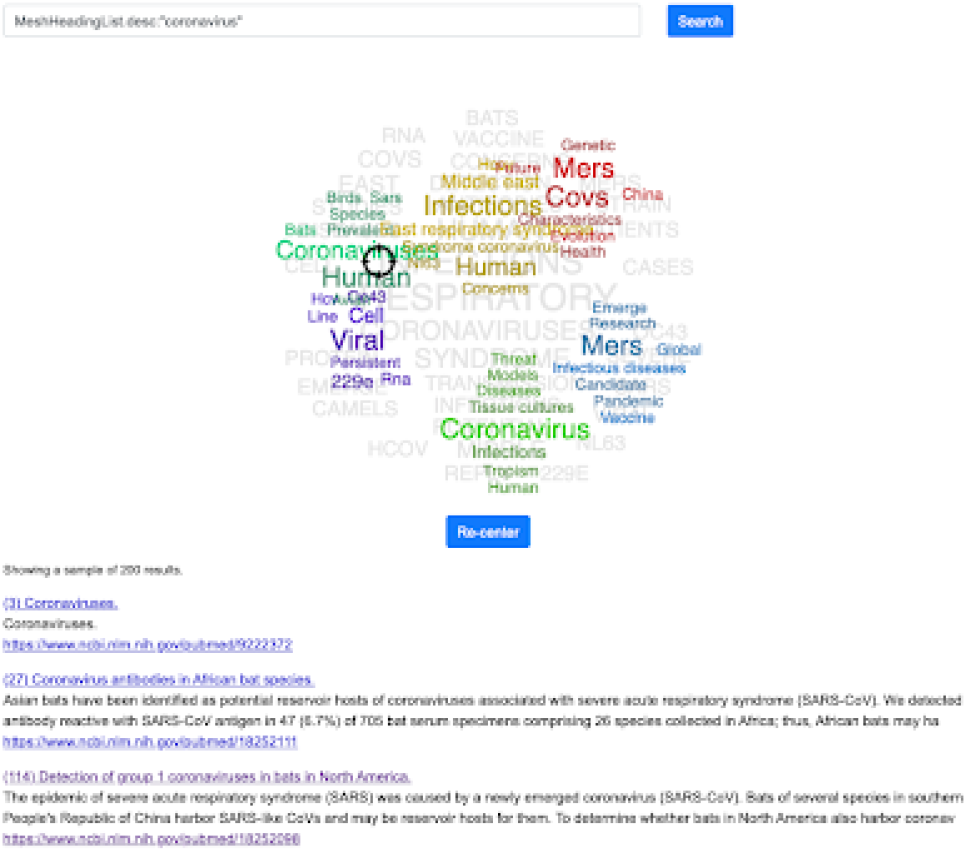
The application of the MEDijs framework to the exploration of health news.

## 6 APPLYING MEDIJS TO AVOID BIAS IN HEALTH NEWS

The appropriate annotation of news is an important value of this technology and is addressed in particular by the integration of the MeSH classifier with the Event Registry system [9] that collects and analyses real-time over 100 thousand news articles daily, offering the user meaningful data visualisation modules to explore their topics of interest. This integration permits health professionals to use the MeSH Headings, to enrich their workflow and enable them to ask the right questions also in the context of worldwide news.

The integration of the MeSH classifier with the worldwide news explorer Event Registry allows us to use MeSH classes in the queries over worldwide news promoting an integrated health news monitoring [6] and trying to avoid bias in this context [7]. An obvious limitation, as earlier mentioned, is a fact that the annotation is only available for news written in the English language, being the unique language in MEDLINE.

The integration of the MeSH classifier with a news engine like Event Registry brings the monitoring of health news to a whole different level. It is leveraging the experience that health professionals have on information tools - MEDLINE and MeSH Headings - that they use frequently on their reviews through, e.g., the PubMed search engine, and builds over these in a smooth incremental manner. In this integration, the health professional can use the MeSH Headings directly at the search of the topic of interest, and those classes will be used to search over the metadata of the previously classified news articles.

In the example of Figure 7 the reader can see a screenshot of the integration of the MeSH Classifier with an example of a news explorer dashboard in utilization, showcasing the dropdown menu where to select the MeSH classes in the news search. This allows also for analytics based on the MeSH classes identified as subtopics of the search query on the input data (see 8.

**Figure 7:**
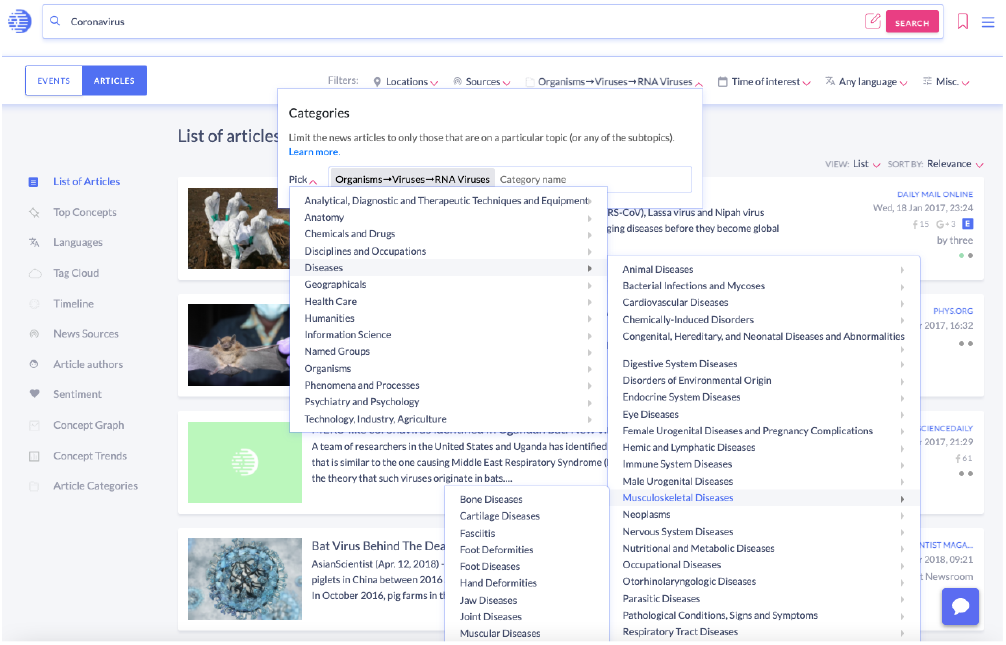
MEDijs allowing a news engine to use MeSH headings as search terms.

**Figure 8:**
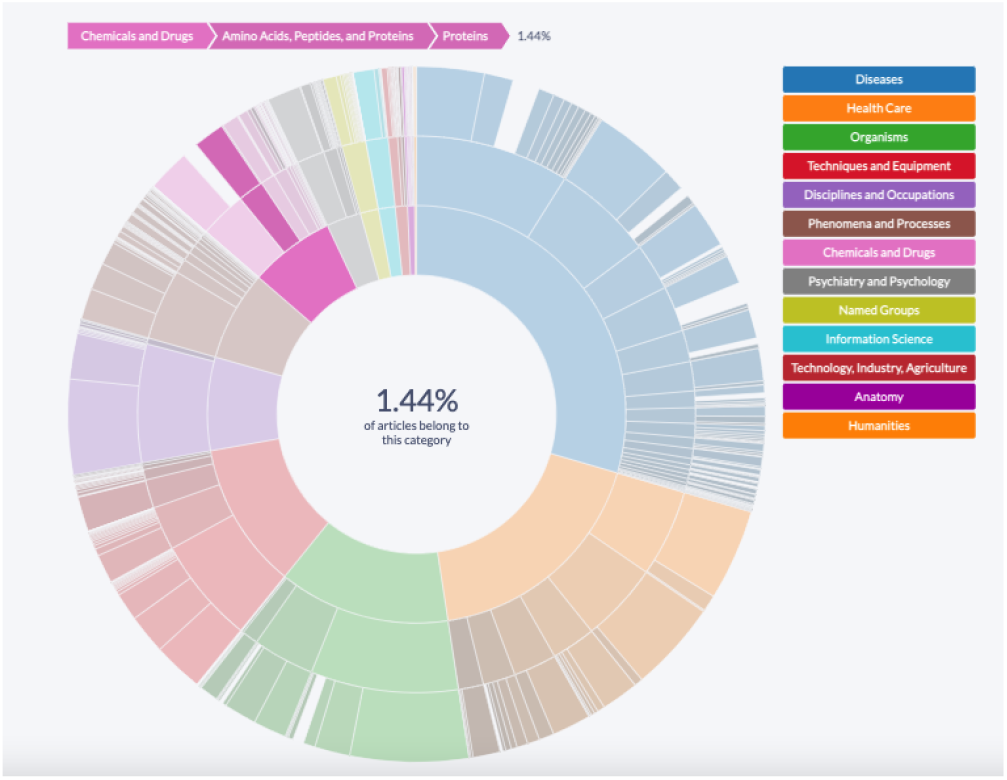
An example of the news data analytics based on the MEDijs framework showing the MeSH-based categories of a query.

Health media bias is a universal concern, particularly in the lessons learned from the COVID-19 crisis in the widespread misinformation. Despite the fact that newspapers and reporters or journalists are to provide the readers with impartial, objective, unbiased and reliable information, the reality is somehow different. Every news story has a potential to be biased, and to be influenced by the attitudes, cultural background, political and economic views of the journalists and editors. Added to this, the infodemics spreads over blogs and social media with a severe impact. In that, some approaches can be taken allowing data exploration and can help balancing the information in worldwide media coverage. The MEDijs in the context of health news, as discussed above, is a central piece for this aim and can provide good ways to explore further what is being talked about. In particular, some of the visualization modules generated by the enriched metadata can help us to explore what is the news coverage of a certain health related measure feeding the general population awareness.

## 7 APPLYING MEDIJS TO GENERATE RECOMMENDATIONS ON MEDICAL REPORTS

The variety of medical reports that can be useful to treat a patient, in the context of healthcare, or to monitor the health of the population, in the context of public health, vary a lot based on their usage and ownership, but also on many other factors including their confidentiality level, Some documents are meant to be made public, others are internal and some are even personal. These differences represent many challenges in the data preprocessing level already, and can impact the meaningfulness of the results of their analysis. The MESH classifier discussed above allows for the integration with third party technologies through a REST API to classify any snippet of health-related text, and thus to the textual content of any medical report or electronic health record. The accuracy of the classification much depends on the amount of data provided to the system, and thus we expect that the usage of it to enrich the metadata of the ingested datasets needs to be used with larger content than those of a few lines of comment to ensure usefulness.

Thus, the performance might me better in summary documents rather than in raw electronic records.

With a large enough amount of historical data, a real-time recommendation system can be established based on the MEDijs framework. The text mining-based recommendation systems are known [4] and can be of great usefulness in the support for decision-making in healthcare [1].

## 8 CONCLUSIONS AND FUTURE WORK

In this work we show the powerful impact of the MEDijs framework in the context of the information retrieval from health-related text through the usage of the controlled vocabulary MeSH and learning algorithms for the text classification based on the MEDLINE dataset. Moreover, we have shown the practical utilization of the interactive data visualisation enabled by the text-mining and machine learning algorithms in use. This shows to be very useful in the exploration of health-related news and, e.g., in avoiding bias in that context. In the future work we will be exploring the potential of recommendation systems based on the MEDijs technology to promote insightful and meaningful suggestions from the input health documents.

## Data Availability

All data indirectly related to this article in the context of the described MeSH classifier can be found at the MIDAS hand-annotated news articles dataset, Zenodo (2020), 10.5281/zenodo.4034281

https://doi.org/10.5281/zenodo.4034281

## ACKNOWLEDGMENTS

The first two authors have been supported by the European Union research fund ‘Big Data Supporting Public Health Policies’, under GA No. 727721.

